# Alpha rhythm-guided repetitive transcranial magnetic stimulation effect on the alpha rhythms of children with autism spectrum disorder – an exploratory EEG study

**DOI:** 10.1101/2025.09.09.25335295

**Authors:** Uchenna Ezedinma, Evan Jones, Alexander Ring, Scott Burgess, Jyoti Singh, Andrew Ladhams, Gary Campell, Shauna Fjaagesund, Piotr Swierkowski, Alexandra Metse, Terri Downer, Florin Oprescu

**Affiliations:** University of the Sunshine Coast; Brain Treatment Centre Australia; Wave Neuroscience inc; Queensland Children’s Lung and Sleep Specialists, Australia; General Practice Unit, Faculty of Medicine, University of Queensland, Herston, Australia; The Thompson Institute, University of the Sunshine Coast, Birtinya, Australia; Health Hub Doctors Morayfield, Australia; School of Psychological Science, University of Newcastle

**Keywords:** Autism spectrum disorder, alpha frequency, repetitive transcranial magnetic stimulation, Electroencephalogram, Children, α-rTMS

## Abstract

**Background:** Quantitative electroencephalogram (qEEG) studies exploring alpha rhythm-guided transcranial magnetic stimulation (α-rTMS) effect on the individual alpha frequency (IAF) of children with autism spectrum disorder (ASD) are sparse.

**Method:** This qEEG study explored the IAF of twenty children (6-12 years old; 16 males) with ASD who were randomly assigned (1:1) to a treatment group (TG) or a waitlist control group (WLCG). The TG received ten α-rTMS sessions over two weeks, while the WLCG acted as control for that period. Next, the WLCG received ten α-rTMS over two weeks. All study participants were followed for one and four months post-study.

**Results:** Following α-rTMS, the TG showed a significant increase in mean frontal region IAF (M =9.093 to 9.351Hz) compared to the WLCG (*F* (1,18) = 6.440, *p = 0.021*, ES = 0.263). When the WLCG received α-rTMS, the mean frontal region IAF increased (M = 8.33 to 8.78Hz) but was insignificant (*F* (2,18) = 2.720, *p = 0.120*, ES = 0.232). The increased regional IAF persisted at one and four months post-α-rTMS follow-up across groups. Additionally, the mean difference between the posterior and frontal region IAF consistently decreased, suggesting improved long-range functional connectivities.

**Conclusion:** This qEEG study presents provisional evidence demonstrating α-rTMS increases IAF within the frontal region of children with ASD and potentially persists for one to four months. Future robust studies using larger sample sizes and more dimensions of electroencephalographic analysis are warranted.

## Introduction

Autism spectrum disorder (ASD) is an increasingly prevalent and complex neurodevelopmental disorder marked by core features of communication and social skill impairments and repetitive behavioural patterns [1]. The global prevalence estimate of children with ASD is documented to be 0.6% [2], with variations ranging between 0.5 – 2.8% across different study samples in Africa, Asia, Australia, Europe and North America [1, 2]. Its increasing prevalence is attributed to greater awareness and early diagnosis [3, 4]. Despite this prevalence, its diagnosis remains behaviourally and not biologically defined [5]. Recent studies have presented electroencephalographic biomarkers, such as alpha rhythm, as a potential marker for biologically defining ASD (For a full review, see Neo et al., [6]). Therefore, as this biomarker may represent a component in the diagnoses of ASD and comorbidities [7], changes to this biomarker following an intervention, such as neuromodulation, is a significant finding [8].

An electroencephalogram (EEG) is widely used to quantitatively measure neural signalling within the neocortex. Its most common signal frequencies are defined into discrete delta, theta, alpha, beta and gamma rhythms. The alpha rhythm originates from the infragranular layers and propagates via layer V pyramidal cells within the cortex. Alpha rhythm is the most prominent eyes-closed, awake and relaxed EEG frequency [9] and has been described to occur within 6-12Hz [9, 10], 7-13Hz [11], 8-12Hz [12], 8-12.9Hz [13], 8-13Hz [14, 15] or 8-16Hz [5]. Nevertheless, its *power, synchrony* and *connectivity* are closely linked to several impaired features, such as cognitive performance, working memory, attention, and social cognition in individuals with ASD [8, 16]. These impaired features are impacted by comorbid sleep difficulties [17, 18], suggesting poorer alpha rhythm characteristics. Thus, core ASD features and sleep comorbidities could be biologically represented by alpha rhythm characteristics.

An individual alpha frequency (IAF) typically increases with age during childhood [6, 9] but is stunted in children with ASD [19, 20]. In age-matched control studies, alpha rhythm was decreased in children with ASD compared to typically developing children [9, 12]. Within the ASD group, alpha rhythm is strongly correlated with cognitive features compared to chronological age [9]. Further, the EEG spectral profile of children with ASD compared to controls shows a U-shaped pattern, i.e., excessive power in delta, theta, beta, and gamma rhythms, with reduced relative alpha rhythm *power* [6, 10, 12]. This diminished alpha *power* also bears decreased long-range functional *connectivity*, especially between the frontal and parietal cortices [13, 21-23]. The long-range functional *connectivity* is determined by the degree of *synchrony* between two regional signals within the EEG band, i.e., the coherence of the underlying brain region [16].

Interestingly, these alpha rhythm characteristics have been experimentally explored in personalising intervention [15, 24] and measuring outcomes such as repetitive transcranial magnetic stimulation [13, 14]. Repetitive transcranial magnetic stimulation (rTMS) is a neuromodulatory technique where an electromagnetically induced field is delivered repetitively to single or multiple cortical sites to alter neuronal activities [25]. rTMS has been shown to induce frequency changes in EEG [26]. In human studies, low frequency (i.e, <5Hz) rTMS causes a long-term suppression of brain excitability by mechanisms related to depression, while high frequency (i.e., >5Hz) rTMS causes a long-term facilitation of brain excitability via mechanisms associated potentiation [27]. While low-frequency rTMS lasts at least 60 minutes, depending on the level of motor cortex excitability, high frequency rTMS can persist up to 90 minutes, depending on the intensity [28, 29].

In personalised rTMS modality, the stimulation frequency could be algorithmically derived from the individual alpha frequency (IAF) and used to guide an attempted restoration of typical alpha rhythm characteristics within underlying cortices of children with ASD [15, 24]. Such modality have been theorised to augment therapeutic response [30], potentially due to its entrainment effects [31]. The algorithmically derived IAF is informed by an age appropriate and typical posterior-to-frontal alpha frequency gradient; that also relies on alpha rhythm generated within the visual cortex of the posterior region [32, 33]. Although atypical alpha rhythms may not be regional-specific in ASD [12, 34], specific regions have been frequently targeted due to their close link with core ASD features, such as the frontal and parietal cortices [35] or based on findings from quantitative EEG studies [34, 36, 37]. However, there is sparse controlled EEG studies on alpha rhythm guided rTMS (α-rTMS) effect on alpha rhythms within the frontal and parietal cortices of children with ASD.

This randomised controlled quantitative EEG study explored the α-rTMS effect on frontal and posterior region IAF of children with ASD. The study hypotheses were:

- The treatment group would show a significant increase in regional IAF following α-rTMS compared to the waitlist control group (phase 1).
- The waitlist control group would show a significant increase in regional IAF following α-rTMS compared to the waitlist period (phase 2).
- The regional IAF of the treatment and waitlist control groups following α-rTMS would persist at one and four months of follow-up (phase 3).

## Methods

### Study design, setting and procedure

This study explored the quantitative electroencephalography (qEEG) data based on data from a randomised waitlist controlled trial [38]. The study received ethics approval from the Ethics Committee of the University of the Sunshine Coast Australia, in February 2023 with ethics number S221766. It was also registered in the Australian New Zealand Clinical Trials Registry (ANZCTR) with number 12623000757617. The study was conducted between October 2023 and September 2024 at a private outpatient α-rTMS centre in North Brisbane, Australia.

In phase 1, following pre-intervention qEEG (T1), the treatment group (TG) received ten sessions (two weeks) of α-rTMS, while the waitlist control group (WLCG) acted as control (waitlist period). Post-intervention qEEG was completed by the groups (T2). In phase 2, following post-waitlist qEEG (T2), the WLCG received the intervention and immediately had waitlist-post-intervention qEEG (T3). In phase 3, both groups were followed up and completed qEEGs at one (T4) and four (T5) months post-study (Figure 1).

**Figure 1:**
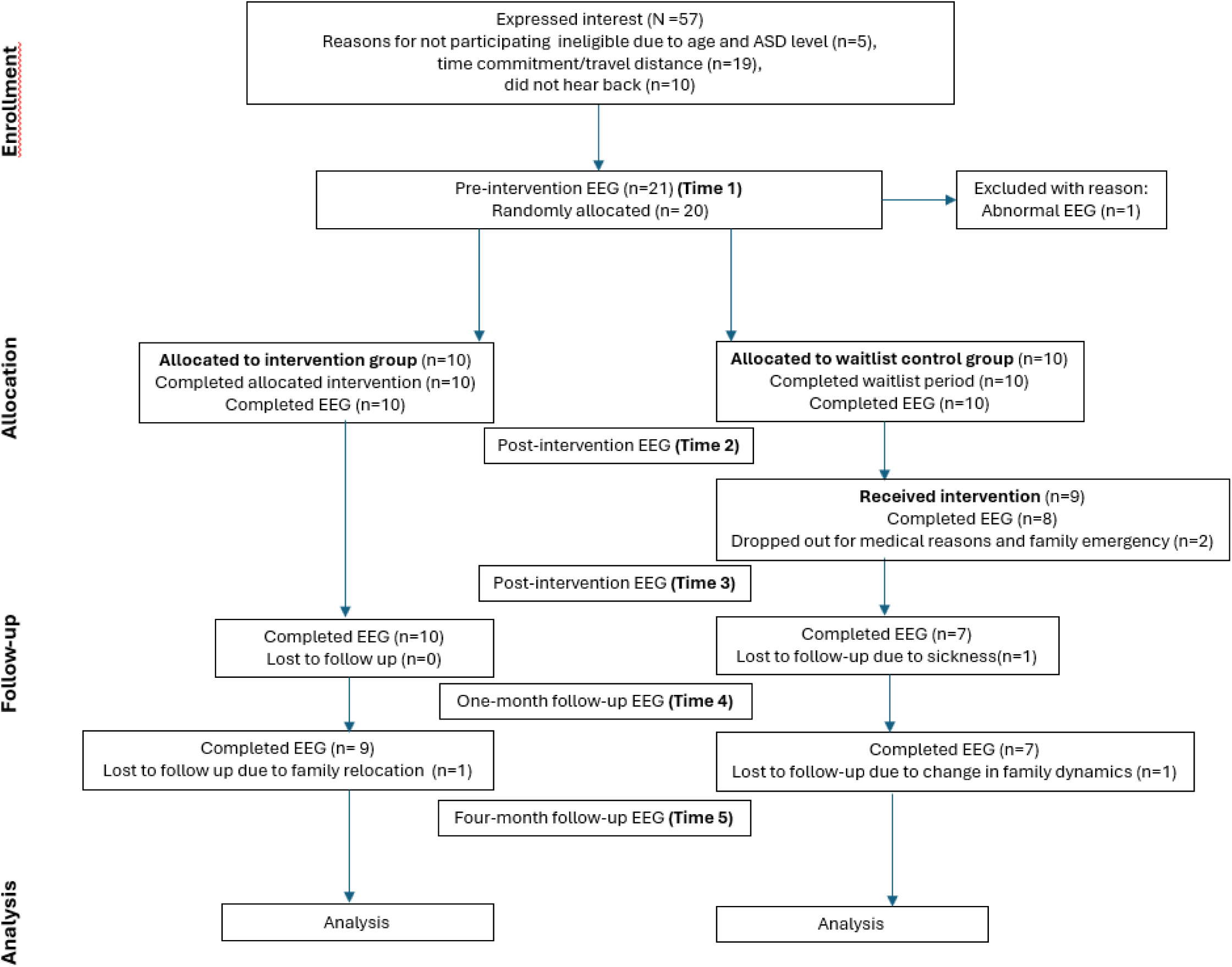
Participant’s flow through the study.

**Figure 2a:**
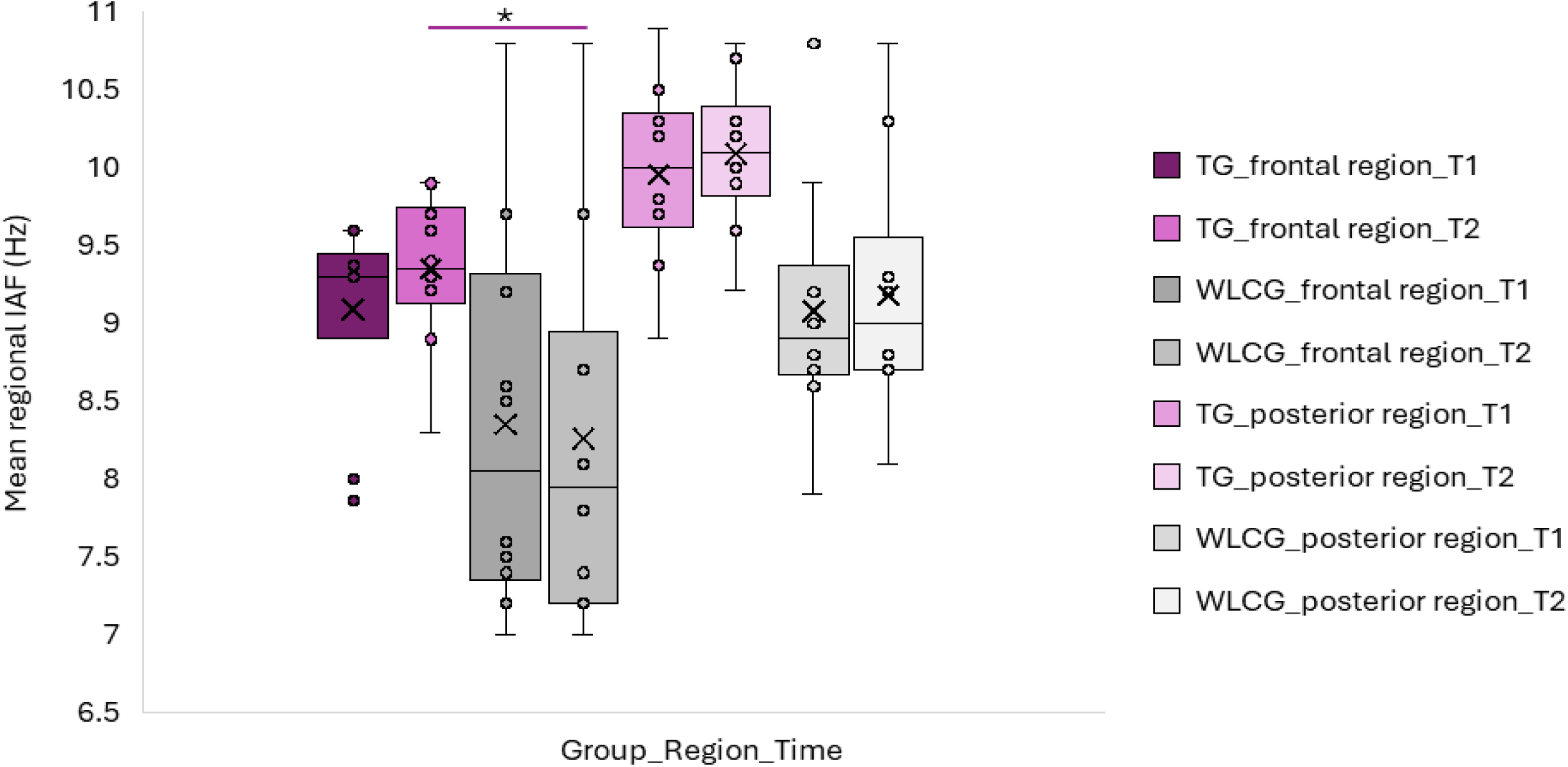
Comparing mean regional IAF between frontal and posterior regions for study phase 1.

**Figure 2b:**
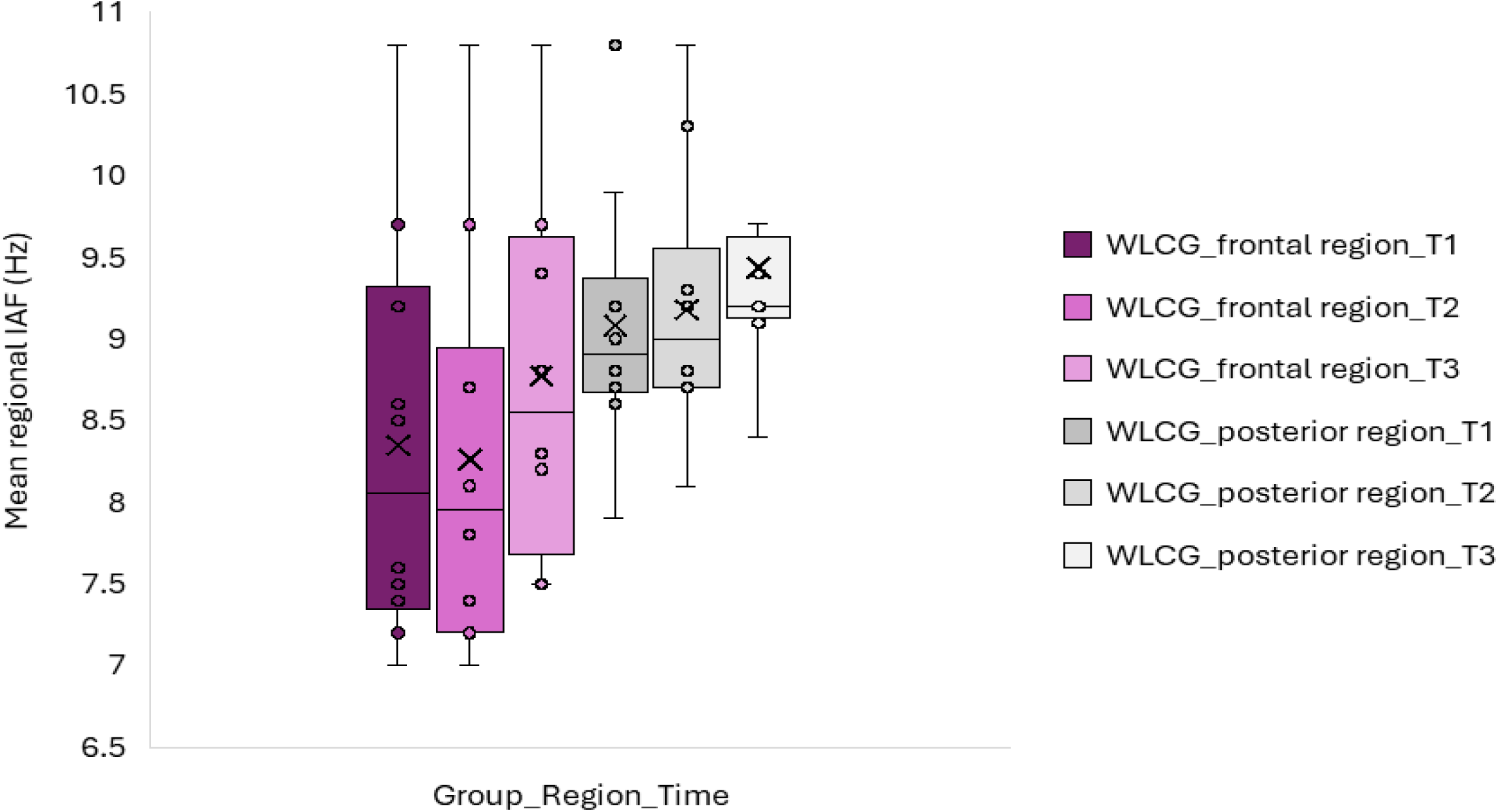
Comparing mean regional IAF between frontal and posterior regions for study phase 2.

### Participants

The participants were recruited from the community via advertisements on relevant institutions’ notice boards, health consumer web pages and social media support groups that targeted primary caregivers of children with ASD. The relevant study inclusion criteria were children (6-12 years old) with a diagnosis of ASD (level 2) based on the Diagnostic and Statistical Manual of Mental Disorders, Fifth Edition, by a paediatrician. This criterion was chosen to ensure sample homogeneity and due to the lack of rTMS studies within this age group [35], who experience a high prevalence of ASD [1, 39]. The study excluded children with (i) qEEGs showing spike and/or slow wave rhythm at pre-intervention (T1). A total of 20 children were randomly assigned to either the TG or WLCG. Participants’ demographic parameters, such as age, gender, and comorbidities, were collected at baseline (Table 1).

**Table 1:** Participant’s demographic characteristics and baseline assessment.

| Baseline characteristics | TG (n=10) | WLCG(n=10) | Comparison |
| --- | --- | --- | --- |
| *Demographics* |  |  |  |
| Mean age (SD)/range | 9 (1.49)/ 7-12 | 9.2 (1.68)/ 6-11 | $p=0.78$ |
| Boys (n) | 7 (70%) | 9 (90%) |  |
| Girls (n) | 3 (30%) | 1 (10%) |  |
| Years since ASD diagnosis (SD)/range | 2.9 (1.79)/1-6 | 3 (1.82)/1-6 | $p=0.90$ |
| *Other comorbidities* |  |  |  |
| Attention Deficit Hyperactivity Disorder | 7 | 7 |  |
| Anxiety (General anxiety disorder, social anxiety) | 6 | 4 |  |
| Dyslexia/ Dyscalculia/ Dysgraphia/sensory disorder | 3 | 2 |  |
| Sleep difficulties | 10 | 10 |  |
| *Existing medications* |  |  |  |
| α-2a adrenergic agonist (clonidine, guanfacine) | 5 | 2 | |
| Melatonin | 4 | 3 |  |
| Stimulant (Lisdexamfetamine, methylphenidate) | 6 | 5 |  |
| *Existing allied health therapies* |  |  |  |
| Counsellor/psychotherapy/psychology | 8 | 4 |  |
| Exercise physiology/physiotherapy | 4 | 1 |  |
| Occupational therapy | 10 | 5 |  |
| Speech therapy | 4 | 3 |  |
| *Baseline regional alpha rhythms* |  |  |  |
| Frontal region | 9.093 | 8.35 | $p=0.050$ |
| Posterior region | 9.957 | 9.08 | $p=0.005$ |

### Intervention

The alpha rhythm (8-13Hz) guided repetitive transcranial magnetic stimulation (α-rTMS) used the proprietary algorithm, commercially known as magnetic EEG-EKG resonant therapy (MeRT), developed by Wave Neuroscience LLC. Briefly, the personalised stimulation frequency was processed using the proprietary algorithm that calculated and identified the participant’s alpha rhythm (8-13Hz range) within the posterior-occipital region of interest (ROI) consisting of P3, P4, Pz, O1, and O2 electrodes. This algorithm has been used in studies involving individuals with posttraumatic stress disorder and comorbid traumatic brain injury or depression [40, 41] but not in individuals with ASD.

This study targeted midline frontopolar and midline parietal cortices. These cortices are part of the six main areas of dysfunction in autism that can be identified using a quantitative electroencephalogram [37]. The fronto-parietal regions also show reduced alpha rhythm power and diminished long-range functional connectivity [21-23]. Both cortices have also been targeted in other α-rTMS studies on children with ASD [15, 42]. The midline frontopolar and midline parietal cortices were located as FPz and Pz, respectively, using the standardised EEG 10-20 positioning [43].

The stimulator output was set at 50% and 40% at the FPz and Pz, respectively. Several high-frequency rTMS studies have used a stimulation intensity ranging from 25-60% [15, 42, 44][21, 45]. Compared to standard rTMS, a reduced intensity is used in α-rTMS, given it is considered to improve outcomes owing to the induction of beneficial forms of neuroplasticity [42]. More so, a reduced stimulation intensity below the motor threshold has been employed in rTMS studies as a safety precaution against seizure risks [46, 47]. The output intensities were also based on the standard procedure of the private outpatient rTMS.

In summary, the α-rTMS protocol was five seconds of stimulation (8.0-13.0Hz range), delivered first to the Pz site and then to the Fpz site, with 28-second intervals between 32 trains and at 40% and 50% output intensity for each site, respectively. The α-rTMS was delivered by a trained technician during consecutive business days (Monday to Friday) on a CF-B65 butterfly coil and Magpro R30 TMS stimulator (Magventure Inc, Denmark). The rTMS coil orientation perpendicular to the FPz and Pz midline [43]. One α-rTMS session is approximately 40 minutes.

### Outcome measure

The primary outcome of this study, regional individual alpha frequency (IAF), was measured on a quantitative electroencephalogram (qEEG). Each participant underwent a ten-minute eyes-closed, awake and relaxed qEEG on a Deymed Truscan EEG system and fitted 21-lead FlexiCAP with standardised 10-20 positioning. qEEG recordings from 38 seconds to 10 minutes are suited to aid compliance in children with ASD and have also been shown to yield reliable and valid data [9, 12-15, 24]. The research assistant (GC) administering the qEEG and the scientist (AR) analysing the data were blinded to the groups.

The EEGs were collected during a 10-minute-eyes-closed awake state with reference electrodes attached to the patient’s earlobes. A linked ear recording reference was used [48]. As per international standards, electrode impedance resistance did not exceed 10 kΩ [49] to ensure electrode connectivity. EEGs were pre-processed with the AutoReject python package, an automated algorithm for rejecting bad EEG epochs [50, 51].

A MNE python package was used to generate a power spectrum density plot as per Welch’s method, which turns time-domain EEG activity into a single spectral frequency plot for each electrode and provides the power of each measured frequency in the EEG and electrodes were averaged in three regions of interest (ROI) including frontal (Fz, F3, F4), central (Cz, C3, C4) and posterior (Pz, P3, P4, O1, O2) leads [52]. The regional individual alpha rhythm (IAF) is the frequency at which the maximum power occurs within the regions of interest. The mean difference between the frontal and posterior regional IAF was used to describe the degree of synchronisation, i.e., long-range functional connectivity.

### Statistical analysis

Analyses were conducted on IBM SPSS, version 29.0. Demographic and baseline data were summarised using standard descriptive and Student *t-*test statistics. The normality of the data was investigated using the Shapiro-Wilk Test. Greenhouse-Geisser (GG) estimates were employed when sphericity was violated.

For study hypothesis 1, a mixed model analysis of variance (ANOVA) was used to assess TG (T1 and T2) versus WLCG (T1 and T2) qEEG. Time (T1 and T2) and group (TG and WLCG) were the within and between-subject variables, respectively. Interaction effects (group x time) indicated whether the intervention improved outcomes for the dependent variable.

For study hypothesis 2, the WLCG data were analysed using repeated-measure ANOVA to determine the main effects of time across the dependent variable. Time (T1 to T3), within-subject variables, were selected as the repeated effects. The main effect of time indicated whether the intervention improved outcomes for the dependent variable.

For study hypothesis 3, TG post-intervention qEEG (T2), waitlist-post-intervention qEEG (T3), one (T4) and four (T5) months post-study follow-up qEEG data were analysed using a linear mixed model for the effect of time across the dependent variable. Time (T2 to T5), within-subject variables, were selected as the repeated effects.

For study hypotheses 1 and 2, a power analysis was also conducted to quantify the risk of Type II error on G*power 3.1software [53]. The effect size was measured as partial eta-squared (ηp^2^) with a small (≥0.01 to ≤0.06), medium (≥0.07 to ≤0.13) and large (≥0.14) scale [54]. All statistical analyses had significance at *p<0.05*.

For each study hypothesis, further analysis of the number of participants with an increase, no change, or decrease in regional IAF was conducted. Pearson’s correlation between the mean difference in IAF following the intervention and the mean difference in frequency (stimulation frequency - baseline IAF) was analysed to validate the proposed hypothesis.

## Results

Table 1 shows a total of 20 children (mean age ±SD = 9.1 ±1.55), comprising 16 boys and 4 girls diagnosed with autism spectrum disorder (level 2), were recruited for this study. Participants had an average of 3 years following ASD diagnosis. All the participants had reported sleep difficulties. Other comorbidities were attention deficit hyperactivity disorder (n=14, 70%), anxiety (n=10, 50%), and sensory or learning difficulties (n=5, 25%). The most used medications included stimulants (n=11, 55%), α-2a adrenergic agonists (n=7, 35%), and melatonin (n=7, 35%), while frequently accessed allied therapies were occupational (n=15, 75%), counsellor/psychotherapy/psychology (n= 12, 60%), speech (n=7, 35%), and exercise physiology/physiotherapy (n=5, 25%).

At baseline, the mean regional IAF at both the frontal and posterior regions was significantly higher in the TG than in the WLCG. Further analysis showed that two participants in the WLCG had a different U-shaped spectral EEG profile suspected to be linked to their methylenetetrahydrofolate reductase (MTHFR) gene variant status, which has been associated with attenuation in IAF at the posterior and frontal region [55].

As shown in Figure 1, eighty-one (90%) of the possible ninety qEEGs were completed across time points, with nine lost to follow-up, mostly in the WLCG, for reasons unrelated to the study: T3 (WLCG: n=2), T4 (WLCG: n=3) and T5 (TG: n=1, WLCG: n=3). Table 2 shows that the mean difference between frontal and posterior IAF was significant for both groups at each time point. The magnitude of the mean differences also showed a steady decrease across time.

**Table 2:** Comparing the differences in the mean regional alpha rhythm between frontal and posterior regions of each group at different time points.

| Time | Group | Frontal IAF (Hz) |  | Posterior IAF (Hz) |  | Posterior-frontal IAF difference | Comparison (p-value) |
| --- | --- | --- | --- | --- | --- | --- | --- |
|  |  | Mean | Range | Mean | Range |  |  |
| T1 | TG (n=10) | 9.09 | 7.9 – 9.6 | 9.96 | 8.9 – 10.9 | 0.87 | 0.0003 |
|  | WLCG (n=10) | 8.35 | 7.0 - 10.8 | 9.08 | 7.9 – 10.8 | 0.73 | 0.0040 |
| T2 | TG (n=10) | 9.35 | 8.3 – 9.9 | 10.09 | 9.21 – 10.8 | 0.74 | 0.0007 |
|  | WLCG (n=10) | 8.26 | 7.0 - 10.8 | 9.18 | 8.1 – 10.8 | 0.92 | 0.0010 |
| T3 | WLCG (n=8) | 8.78 | 7.5 – 10.8 | 9.44 | 8.4 – 11.3 | 0.66 | 0.0200 |
| T4 | TG (n=10) | 9.50 | 8.7 – 10.3 | 10.10 | 9.2 - 11.3 | 0.60 | 0.0009 |
|  | WLCG (n=7) | 8.99 | 7.8 – 10.6 | 9.51 | 8.8 – 10.6 | 0.52 | 0.0400 |
| T5 | TG (n=9) | 9.71 | 8.7 – 10.3 | 10.18 | 9.5 – 11.3 | 0.47 | 0.0090 |
|  | WLCG (n=7) | 8.79 | 7.8 - 10.1 | 9.43 | 8.2 – 10.6 | 0.64 | 0.0090 |

**Hypothesis 1:**

Table 3a shows that the group x time interaction for the TG mean IAF at the frontal region significantly increased compared to the WLCG (*F* (1,18) = 6.440, *p = 0.0210*, ES = 0.263 The group x time interaction for the TG mean IAF at the posterior region did not significantly change compared to the WLCG (*F* (1,18) = 0.090, *p = 0.7680*, ES = 0.005).

**Table 3a:** Comparing mean regional IAF between frontal and posterior regions for study phase 1.

| Region | TG (n=10) |  | WLCG (n=10) |  | Group*time interaction |  |  |
| --- | --- | --- | --- | --- | --- | --- | --- |
|  | T1 | T2 | T1 | T2 | F (df=1, 18) | p-value | Effect size |
| Frontal | M (SD)<br>9.09 (0.6) | M (SD)<br>9.35 (0.5) | M (SD)<br>8.35 (1.2) | M (SD)<br>8.26 (1.2) | 6.440 | **0.021** | 0.263 |
| Posterior | M (SD)<br>9.96 (0.6) | M (SD)<br>10.10 (0.5) | M (SD)<br>9.08 (0.8) | M (SD)<br>9.18 (0.8) | 0.090 | 0.768 | 0.005 |

The changes in IAF within the frontal and posterior regions (T2 vs T1) were examined for each participant. Five (diff: 0.3 to 0.9Hz), four (diff: 0Hz) and one (diff: -0.16Hz) TG participants compared to two (diff: 0.1-0.3Hz), five (diff: 0Hz) and three (diff: -0.5 to -4Hz) WLCG participants showed increases, no differences and decreases in frontal IAF, respectively. On the other hand, five (diff: 0.1 to 0.7Hz), two (diff: 0Hz), and three (diff: -0.3 to -0.1Hz) TG participants compared to five (diff: 0.1 to 0.4Hz), four (diff: 0Hz), and one (diff: -0.1Hz) WLCG participants showed an increase, no change, and decreased posterior IAF, respectively.

From Table 2, the difference between the mean IAF within the frontal and posterior regions were further examined for the TG and WLCG at T2 vs T1. The posterior-frontal region IAF difference reduced in the TG (M= -0.13Hz) and increased in the WLCG (M= 0.19Hz) at T2 compared to T1. Such decrease in IAF between the frontal and posterior regions for the TG was significant compared to the WLCG (M = 0.06; *p = 0.0300)*.

The stimulation frequency of participants in the TG ranged between 8.99 and 11.2Hz (M= 10.10Hz). Further analysis of the frontal (r=0.76, *p= 0.0005*) and posterior (r=0.02, *p= 0.8740*) regions of the TG showed a positive correlation between the mean difference in IAF (T2 vs T1) versus the mean difference in frequency change (stimulation frequency vs IAF at T1).

**Hypothesis 2:**

For this analysis, two participants with missing data at T3 were excluded. Table 3b shows the mean IAF at the frontal (*F* (2,14) = 2.805, *p = 0.0950*, ES = 0.286) and posterior (*F* (2,14) = 3.684, *p = 0.0860*, ES = 0.345) regions following the intervention at T3, which was not significantly different compared to the end of the waitlist period (T2). With an achieved power of 0.950 and 0.907, the risk of type II error was 0.05 and 0.09, for the frontal and posterior region, respectively.

**Table 3b:** Comparing mean regional IAF between frontal and posterior regions for study phase 2.

| Region | Mean value (n=8) |  |  | Effect of time |  |  |
| --- | --- | --- | --- | --- | --- | --- |
|  | T1 | T2 | T3 | F (df=2, 14) | p-value | Effect size |
| Frontal | M (SD)<br>8.46 (1.3) | M (SD)<br>8.33 (1.3) | M (SD)<br>8.78 (1.1) | 2.805 | 0.095 | 0.286 |
| Posterior | M (SD)<br>9.18 (0.9) | M (SD)<br>9.30 (0.9) | M (SD)<br>9.44 (0.8) | 3.684 | 0.086 | 0.345 |

The changes in IAF within the frontal and posterior regions (T3 vs T2) were examined for each participant. Six (diff: 0.1 to 1.8Hz) and two (diff: 0Hz) participants showed increases and no differences in frontal IAF at T3, respectively. On the other hand, five (diff: 0.2 to 0.5Hz), one (diff: 0Hz), and two (diff: -0.6 to -0.1Hz) participants showed increases, no difference and decreases in posterior IAF at T3, respectively.

From Table 2, the differences between the mean IAF within the frontal and posterior regions was further examined at T2 vs T3. The posterior-frontal region IAF difference was reduced at T3 (M= - 0.3Hz, p= 0.095) compared to T2.

The stimulation frequency of participants in the WLCG ranged between 8.4 and 11.0Hz (M= 9.54Hz). Further analysis of the frontal (r=0.51, *p= 0.0210)* and posterior (r=0.32, *p= 0.0990*) regions of the WLCG showed a positive correlation between the mean difference in IAF (T3 vs T2) versus the mean difference in frequency change (stimulation frequency vs IAF at T2).

**Hypothesis 3:**

For this analysis, five participants (TG = 1, WLCG = 4) with missing data at T3, T4 and T5 were excluded. Table 3c shows that for both groups and at the frontal and posterior regions, the mean IAF increased at one (T4) and (T5) months post-study follow-ups compared to T2/T3. Only the difference in the frontal region IAF between T5 vs T2 of the TG was significant (M=0.31, *p= 0.0116)*. For both groups and at the frontal and posterior regions, the mean difference in IAF followed a similar trend of an increase between T4 vs T2/T3 and T5 vs T2/T3, but decreased between T5 vs T4.

**Table 3c:**
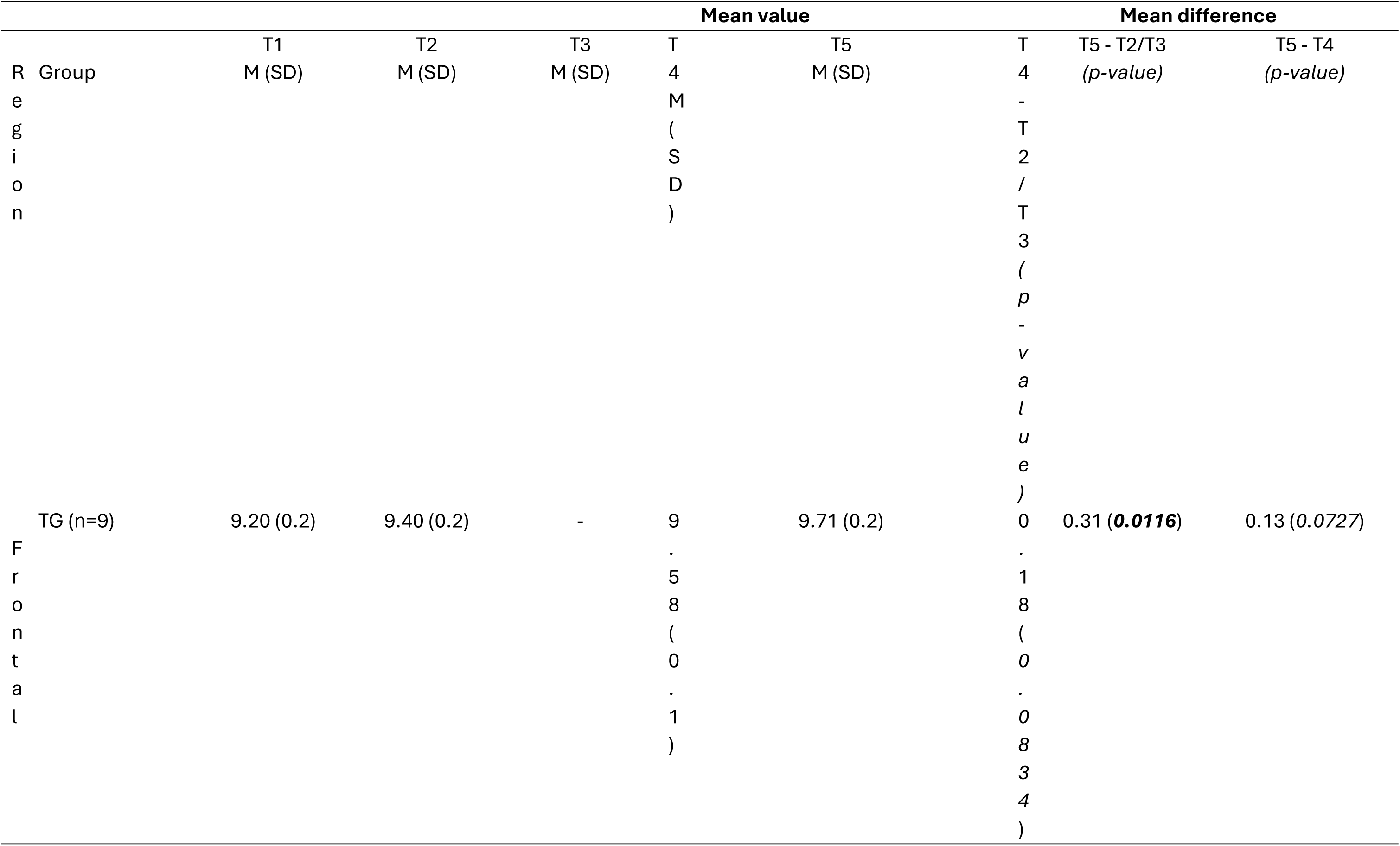
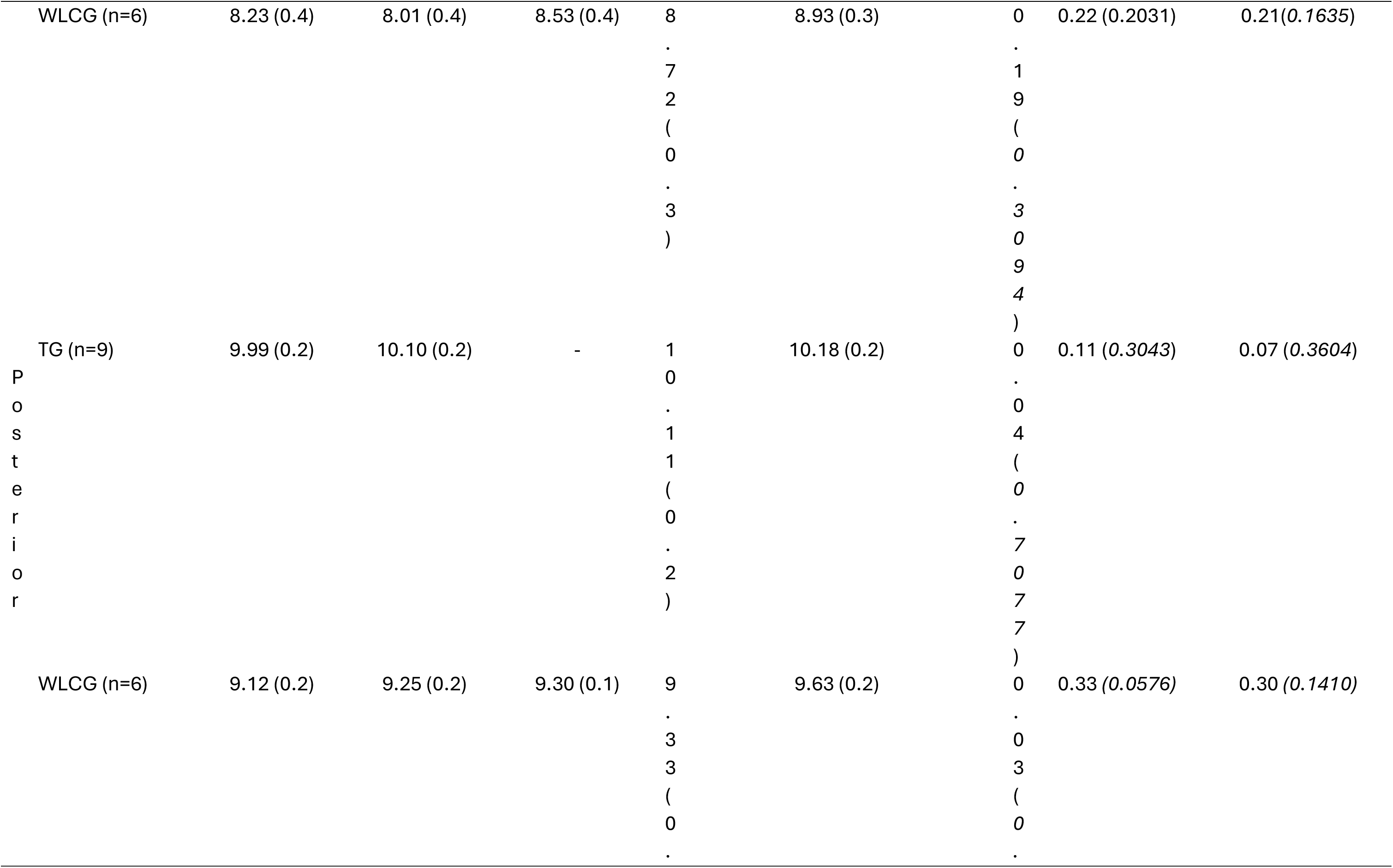
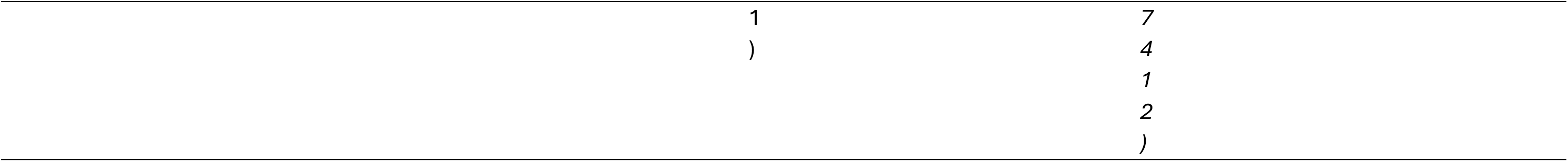
Comparing mean regional IAF between frontal and posterior regions for study phase 3.

The changes in IAF within the frontal and posterior regions (T4 vs T2/T3 and T5 vs T2/T3) were examined for each participant. For the TG at T4 vs T2, seven (diff: 0.03 to 1.0Hz) and two (diff: 0Hz) participants showed increases and no differences in frontal region IAF, respectively, while six (diff: 0.1 to 0.4Hz), one (diff: 0Hz) and two (diff: -0.3 to -0.2Hz) participants showed increases, no difference and decreases in posterior region IAF, respectively. At T5 vs T4, six (diff: 0.1 to 0.4Hz), one (diff: 0Hz) and two (diff: -0.1Hz) TG participants showed increases, no difference and decreases in frontal region IAF, respectively, while at the posterior region IAF, four (diff: 0.1 to 0.4Hz), three (diff: 0Hz) and two (diff: -0.2Hz) participants showed increases, no differences and decreases, respectively.

For the WLCG at T4 vs T3, three (diff: 0.2 to 0.9Hz), one (diff: 0Hz) and two (diff: -0.2 to -0.1Hz) participants showed increases, no difference and decreases in frontal region IAF, respectively, while three (diff: 0.2Hz), two (diff: 0Hz) and one (diff: -0.1Hz) participants showed increases, no differences and decrease in posterior region IAF, respectively. At T5 vs T4, three (diff: 0.1 to 0.8Hz) and three (diff: 0Hz) WLCG participants showed increases and no difference in posterior region IAF, respectively, while at the posterior region IAF, four (diff: 0.2 to 0.9Hz), one (diff: 0Hz) and one (diff: -0.2Hz) participant showed increases, no differences and decreases, respectively.

Further analysis shows that the posterior-frontal region differences in the TG were reduced at T4 and T5 compared to T2 (T4 vs T2: M = -0.13, *p = 0.3796* and T5 vs T4: M = -0.2, *p= 0.1922).* The WLCG showed T4 vs T3 reduced (M = -0.15, *p = 0.5557*) while T5 vs T4 increased (M = 0.08, *p = 0.4742*) in posterior-frontal region differences.

## Discussion

This study demonstrated that ten sessions of alpha rhythm guided repetitive transcranial magnetic stimulation (α-rTMS) significantly increased individual alpha frequency (IAF) within the frontal brain region of children with autism spectrum disorder (ASD) as measured on quantitative electroencephalogram (EEG).

In study phase 1, α-rTMS significantly increased the IAF within the frontal region of participants in the treatment group compared to the control group. This finding may suggest that frontal regional IAF are more responsive to frequency-modifying effects from α-rTMS than within the posterior region. The lack of a significant α-rTMS effect on the posterior region IAF may be due to the stimulation frequency being determined from the posterior electrodes and its proximity to the posterior region rather than the frontal region’s IAF. That is, with the mean frontal region IAF of 9.09Hz and the mean posterior region IAF of 9.96Hz at T1, the mean stimulation frequency of 10.10Hz would not necessarily impact a significant change on the posterior compared to the frontal region, given that the absolute difference between stimulation frequency and local alpha frequency is much higher for the latter than the former.

Alternatively, it could be that the posterior region mean IAF did not increase beyond 10.10Hz, potentially due to the entraining effect from the mean stimulation frequency of 10.10Hz [31]. Such electroencephalographic changes in alpha rhythm following α-rTMS may also be supported by a positive correlation between the mean change in regional IAF and the difference between the stimulation frequency and baseline IAF. For instance, a 0.5Hz change in a region’s IAF following α-rTMS positively correlated with a 0.5Hz difference between the stimulation frequency and baseline IAF. Furthermore, as individuals with autism bear diminished long-range functional connectivity [13, 21-23], the reduction in the difference between the mean posterior and frontal region IAF suggests improved connectivity and coherence of the underlying brain regions [16].

However, it must be noted that the significant improvement in frontal region IAF and long-range functional connectivity in the TG compared to the WLCG may be due to disparities at baseline. The IAF at the frontal and posterior regions was significantly higher in the TG than in the WLCG. This finding may suggest that regional IAF for the WLCG was delayed considerably for their age. In fact, for a mean age of 9.2 years, the frontal region IAF (8.35Hz) in the WLCG is lower compared to another waitlist-controlled rTMS study of 16 low-functioning autistic children with a mean age of 7.8 years and corresponding mean frontal IAF of ∼9.39Hz at baseline [14]. While in another EEG study, an IAF of 8.89Hz was related to 7-year-old typically developing children [56]. Age-related factors, such as the age range within the group, may also have impacted the IAF in the WLCG. In this study, the TG (7-12 years old) had a higher age range than the WLCG (6-11 years old). Edgar et al. [57] reported that higher IAF were observed in a study on older autistic children (6 -17 years old) compared to younger autistic children (2-12years old).

There is also the potential impact of gender differences. For instance, the ratio of males to females between both groups may have determined the differences in IAF values [57]. The interaction between gender and comorbidities such as ADHD may relate to IAF values via the hypothesised role of sleep on alpha rhythm [58]. To illustrate this, sleep studies on children with ADHD have reported longer total sleep time in the female than in the male participants [59, 60]. With each group having equal ADHD comorbidities but a higher number of females than males in the TG than WLCG, it could be suggested that a longer total sleep time in the former underpinned the higher IAF than the latter. As participants in the WLCG received usual care during the waiting period, it is also possible that a change in usual care, such as medication [16] and allied therapy use during the period, could explain the worsening of frontal region IAF and long-range connectivity. Such diverse disparities at baseline may have confounded the differences in alpha rhythm characteristics between the TG and WLCG.

In study phase 2, the administration of α-rTMS to the WLCG was expected to further shed more light on the effect of the intervention on regional IAF. Following α-rTMS at T3, the mean IAF in the frontal and posterior regions improved, albeit not significantly, compared to the waiting period at T2. Unlike participants in the TG, the lack of a significant increase in the frontal region IAF may be due to (i) participants’ drop-out impacting the analysis (ii) two participants had a distinct EEG pattern - prominent bilateral parieto-temporally generated 4.5 Hz rhythm – potentially linked to their methylenetetrahydrofolate reductase (MTHFR) gene variant status. A recent report on 29 children (2-12 years old) with ASD suggested that the presence of the distinct EEG pattern may significantly impact an otherwise attenuated IAF, especially within the frontal region [55]. Such an impact of the distinct EEG pattern on regional IAF may also have influenced a lower stimulation frequency (M= 9.54Hz) for participants in the WLCG compared to the age-matched participants in the TG.

Therefore, the lack of significant change in frontal region IAF within WLCG should be interpreted within the context of the distinct EEG pattern. Notwithstanding, compared to the waiting period, the frontal region IAF of most participants increased while the posterior region IAF remained the same. Like in the TG, this finding may suggest that frontal regional IAF are more responsive to frequency-modifying effects from α-rTMS than within the posterior region. Also, the lack of a significant α-rTMS effect on the posterior region IAF may be due to the stimulation frequency being determined from the posterior electrodes and its proximity to the posterior region rather than the frontal region’s IAF. Other similarities with the TG are the positive correlation between the mean change in regional IAF and the difference between the stimulation frequency and baseline IAF. There was also a similar reduction in the mean IAF between the frontal and posterior regions following α-rTMS, suggesting an improvement in long-range functional connectivity.

In study phase 3, it would be suggested that the effects of α-rTMS persisted for up to four months post-intervention, given that the regional IAF values for participants within the TG and WLCG remained above pre-α-rTMS values. In fact, the regional IAF values for both groups at one and four months follow-up consistently increased compared to post-α-rTMS, albeit only the difference between TG’s frontal region IAF at four-month follow-up was significant. Also, the mean difference between the frontal and posterior region IAF for both groups generally demonstrated a steady decrease, suggesting an increase in long-range functional connectivity [13, 21-23]. As the midline of the stimulated frontal and posterior regions bears the highest IAF in children with ASD [20], it can be postulated that α-rTMS of the midline region may have led to the persistent increase in the IAF of study participants for up to four months.

Prior open-label α-rTMS study had reported on the significant increase in the IAF of autistic children (mean age of 6.1 years); however, this is the first study to conduct a post-α-rTMS follow-up. It is worth noting that despite the lack of a control group in this study phase, the increasing IAF may not be due to age [19, 20]. Although a change in IAF of children with ASD is thought to be a function of age ASD [20]. Therefore, this may explain why, within individual analysis, there was heterogeneity in IAF changes in both regions and across study groups. For instance, the difference in age range between both groups may explain why seven and six TG participants, compared to three and three WLCG participants, had an increase in the frontal region IAF at one and four month follow up, respectively. It is still possible that the changes in regional IAF during the post study follow up may be due to medications and allied therapy use, amongst other factors not controlled for.

### Clinical relevance

The α-rTMS effect on alpha rhythms in this study is clinically relevant [8]. Firstly, the study is relevant for the biologically-defined assessment compared to behaviourally-defined assessments within the population [5]. The significant difference in alpha rhythms between the age-matched groups can enable the biological assessment of the child’s development based on cognitive processing rather than chronological age [9]. This biological assessment tool may be more useful in sampling study participants within a research setting.

Secondly, the participant’s regional IAF may be relevant for assessing ASD and other frequently reported comorbidities, such as ADHD and sleep difficulties, in this study [7]. For instance, it’s being reported that children with ASD have widespread abnormal alpha activity across distal regions, which impacts long-range functional connectivity. On the other hand, comorbid ADHD have been consistently found to be associated with frontal region dysfunction [12] while comorbid sleep difficulties have been reported to be closely related to the role of alpha rhythm on arousal [12] and sleep [58].

Thirdly, the increase in regional IAF in this study provides biologically defined evidence of the potential response of children with ASD to the entrainment effect of α-rTMS [31]. Such a change in IAF may be an objective tool for predicting the therapeutic response of participants to α-rTMS. For instance, the increasing alpha rhythm towards 10Hz —the centre of the EEG alpha rhythm – may predict clinical response [30]. Therefore, it could be argued that the increased frontal region IAF within the TG predicted the positive sleep difficulties as reported elsewhere [38].

Finally, this EEG study may help determine the clinical value of algorithms used in developing the protocol for α-rTMS. This position is supported by the fact that the direction and magnitude of rTMS effect depend on stimulation parameters (intensity, frequency, number of sessions, etc) and functional condition of the targeted region [35]. Based on the study outcomes, the stimulation parameters such as the frequency of 8.0-13.0Hz, 40-50% output intensity and the highly sensitive frontal region, may be a therapeutic stimulation parameter within the population.

#### Study Limitations and Strengths

This is the first randomised controlled trial study reporting on the α-rTMS effect on the frontal and posterior IAF of children with ASD. Previous studies used an open-label study design [15, 42]. Thus, this study presents controlled data on the α-rTMS effect on IAF of autistic children. It adds to the body of evidence from a sham study that concluded that α-rTMS might be a valuable therapeutic tool for individuals with cortical dysfunction [11], such as ASD, potentially due to increased alpha rhythm and long-range connectivity (*coherence*) in cortical regions. The recruitment of participants between 6-12 years old and ASD level 2 improved the homogeneity within the sample, thus strengthening the internal validity of the findings within the cohort, a stratified study [61]. More so, the male-to-female ratio of 4:1 is comparable to that of epidemiological studies [62, 63]. This gender distribution suggests that the study population is representative of the broader ASD population. The IAF difference between age-matched randomised groups suggests the value of EEG in personalising rTMS frequency compared to using an age-estimated rTMS frequency [24]. More so, stimulating multiple regions improves the chances of therapeutic response in autistic individuals who bear global alpha rhythm dysfunction [12]. This study is also strengthened by its long-term follow-up to identify the duration of α-rTMS effect on the IAF of children with ASD.

However, this study is limited by its small sample size, the recruitment of participants of a specific age (6-12 years) and ASD diagnosis (level 2) and a short course of α-rTMS sessions. The study recruitment criteria may limit the external validity of its findings within the heterogeneous ASD population. For instance, the finding may not hold for children outside the 6-12 years range as the IAF may vary at different ages [6]. The sample size may limit the accurate estimation of the α-rTMS effect. The potential confounders, such as genetics, medication, sleep difficulties [64] and the time of EEG collection [65] on α-rTMS effect, were not controlled for. The relatively small sample size limited the use of these potential confounders as covariates in the analysis. Ten α-rTMS sessions may not have been sufficient to produce a more durable clinical and lasting outcome as reported in other rTMS studies [13-15, 24].

#### Recommendation

A large sample of pharmacological and therapy-naïve participants across the child age range and ASD levels is required to accurately determine the effects of α-rTMS on the regional alpha rhythms within a randomised sham or placebo-controlled trial design. Such studies may consider accounting for differences in participant alpha rhythms rather than chronological age at baseline [9]. Future studies using a longer course of α-rTMS sessions and utilising EEG measures in connectivity and complexity domains, such as entropy for the alpha rhythm power and other frequency bands, should be considered.

## Conclusion

Electroencephalographic studies show that alpha rhythm and long-range functional connectivities are impaired in children with autism spectrum disorder. This study demonstrates that alpha rhythm-guided repetitive transcranial magnetic stimulation significantly increased the regional alpha rhythm within the frontal brain region of children with ASD, and the effect lasted from one to four months. The posterior-frontal IAF difference decreased over four months following α-rTMS, suggesting an improved long-range functional connectivity. Future robust studies using larger sample sizes and more dimensions of electroencephalographic analysis are warranted.

## Data Availability

Mediated access to non-identifiable study data is available from the corresponding author on written approval from funders to further protect the privacy and sensitivity of the health information of children participants.

